# Immunogenicity and safety of DS-5670d, an mRNA-based COVID-19 vaccine targeting omicron XBB.1.5: Results from a phase 3, randomized, active-controlled study in adults and children aged ≥12 years

**DOI:** 10.1101/2024.11.14.24317321

**Authors:** Ami Kawamoto, Masahiro Hashida, Katsuyasu Ishida, Kei Furihata, Aisaku Ota, Kaori Takahashi, Sachiko Sakakibara, Takashi Nakano, Fumihiko Takeshita

## Abstract

**Background:** Many people worldwide have now acquired immune responses against the severe acute respiratory syndrome-coronavirus-2 (SARS-CoV-2) following previous vaccination and/or infection. As a result, national vaccination programs are now implementing a simplified schedule of single-dose administration and seasonal boosters. In this phase 3 non-inferiority study, we assessed the immunogenicity and safety of a single dose of the lipid nanoparticle-messenger ribonucleic acid vaccine DS-5670d, a monovalent composition for the 2023/24 season, containing an omicron XBB.1.5-derived antigen.

**Methods and Findings:** Adults and children aged ≥12 years were stratified according to their history of both prior SARS-CoV-2 infection plus prior coronavirus disease 2019 (COVID-19) vaccination (subpopulation A), prior infection only (subpopulation B), prior vaccination only (subpopulation C), or no history of either infection or vaccination (subpopulation D), and randomly assigned (1:1) to receive DS-5670d or monovalent BNT162b2 omicron XBB.1.5. In the combined ABC subpopulations (DS-5670d, *n* = 362 vs BNT162b2, *n* = 363), the adjusted geometric mean titer ratio of blood neutralizing activity against SARS-CoV-2 (omicron XBB.1.5) was 1.218 (95% confidence interval [CI], 1.059, 1.401) and the seroresponse rates were 87.3% (DS-5670d) and 82.9% (BNT162b2) with an adjusted difference of 4.5% (95% CI, –0.70, 9.71). Both results exceeded prespecified non-inferiority margins and the study met the primary endpoint. Immunogenicity data in the overall ABCD population also met non-inferiority criteria. There were no apparent differences in immunogenicity according to age or sex, and analyses suggested that even unvaccinated persons achieved an adequate immune response following a single dose of DS-5670d. There were no major differences in the incidence or severity of adverse events between the study vaccination groups.

**Conclusions:** A single dose of DS-5670d was immunogenically non-inferior to BNT162b2 and acceptably safe in persons with or without a history of prior infection and/or vaccination.

**Trial Registration:** Japan Registry of Clinical Trials (jRCT2031230424)

**Author summary:** **Why was this study done?**

- The global emergency response against the coronavirus disease 2019 (COVID-19) pandemic is now being superseded by annual immunization using an updated vaccine composition to reduce the disease burden associated with newly emerging variants.
- We conducted a comparative study of two newly authorized updated vaccine compositions containing the antigen derived from omicron XBB.1.5, which was recommended for the 2023/24 season.
- We intended to assess the immunogenicity and safety of a single dose of DS-5670d compared with BNT162b2 in Japanese adults and children aged ≥12 years.

**What did the researchers do and find?**

- In persons who had received prior vaccination, or who had previously had COVID-19, or both, DS-5670d was non-inferior to BNT162b2 in terms of blood neutralizing activity and seroresponse rates, and immune responses were not influenced by age or sex.
- Among unvaccinated persons, those who had been exposed to SARS-CoV-2 by prior infection achieved an adequate immune response following a single dose of DS-5670d; however, the group who had had no previous exposure at all was too small to properly evaluate.
- There were no major differences in the frequency or severity of adverse events between the study vaccination groups, and there were no DS-5670d-related serious adverse events.

**What do these findings mean?**

- DS-5670d was immunogenically non-inferior to BNT162b2, which has already been widely administered in Japan.
- There were no new critical safety concerns associated with DS-5670d, suggesting that it could be a useful SARS-CoV-2 vaccine option.
- Taken together, the results from this study of DS-5670d and those from previous studies evaluating other compositions of DS-5670, i.e., those containing antigens from different strains of SARS-CoV-2, provide corroborating evidence that the DS-5670 vaccine platform can be applied to produce effective and safe seasonal vaccines against future emerging strains.

## Introduction

Although coronavirus disease 2019 (COVID-19) no longer constitutes a global emergency, it remains an ongoing threat to public health [1]. The emergence of novel escape variants of severe acute respiratory syndrome-coronavirus-2 (SARS-CoV-2), plus waning protective immunity over time, has led to a continuous need for the development, testing and authorization of updated vaccine compositions [2, 3]. The initial roll-out of primary vaccination with monovalent messenger ribonucleic acid (mRNA)-based vaccines such as BNT162b2 (Comirnaty, Pfizer-BioNTech) and mRNA-1273 (Spikevax, Moderna) targeting the original Wuhan strain of SARS-CoV-2 [4] was initially followed by boosters with the same monovalent composition, and then by bivalent boosters targeting both the original strain and omicron BA.4-5 [5, 6]. Immunization programs against COVID-19 have now moved away from emergency use towards seasonal vaccination, to boost protective immune responses against emerging variants of SARS-CoV-2 [1, 7]. In addition, a single-dose regimen is now recommended for most recipients, as either a primary or booster vaccination [8, 9]. For the 2023/24 season, monovalent vaccine compositions containing an antigen from the omicron XBB.1.5 strain were authorized [10], while for 2024/25, vaccines containing an antigen from the JN.1 lineage have been recommended [11, 12].

DS-5670 is a vaccine platform formulation based on mRNA encapsulated in lipid nanoparticles (LNP-mRNA) [13], which can be applied to compositions containing antigens derived from different strains of SARS-CoV-2. To date, several different compositions of DS-5670, either monovalent or bivalent, have been clinically evaluated. DS-5670a, a monovalent vaccine containing an antigen from the original strain, demonstrated a clinically acceptable safety profile and favorable immune responses in adults [14], and was authorized in Japan for use as a booster vaccination. DS-5670a/b, a bivalent vaccine containing antigens from both the original strain and from omicron BA.4-5, was shown to be well-tolerated, induced broad neutralization activity across omicron sub-lineages, and was immunogenetically non-inferior to the bivalent composition of BNT162b2 containing corresponding original and omicron BA.4-5 antigens when administered as a booster vaccination in adults (Daiichi Sankyo Co., Ltd.; data on file). DS-5670a/b also had a manageable safety profile and showed non-inferior immunogenicity to bivalent BNT162b2 when administered as a booster to children aged 5–11 years [15].

In November 2023, a monovalent DS-5670 vaccine containing an antigen derived from omicron XBB.1.5 (DS-5670d; Daichirona^®^, Daiichi Sankyo Co., Ltd.) was approved by the Japanese Ministry for Health, Labour and Welfare as a booster COVID-19 vaccination [16], and it was launched in December as part of the Japanese immunization program for the 2023/24 season. Herein, we report data from a phase 3 non-inferiority study conducted in adults and children aged ≥12 years which aimed to evaluate the safety and immunogenicity of a single dose of DS-5670d in those with or without history of prior infection and/or vaccination.

## Methods

### Participants

This study included adults and children aged ≥12 years enrolled from 15 sites across Japan (**S1 Table**). Written informed consent was obtained from each participant or their legal representative prior to initiation of study procedures. Eligible participants were stratified into 4 subpopulations: (A) those with a history of both SARS-CoV-2 infection and COVID-19 vaccination, (B) those with a history of SARS-CoV-2 infection but without a history of COVID-19 vaccination, (C) those without a history of SARS-CoV-2 infection but with a history of COVID-19 vaccination, or (D) those without any history of SARS-CoV-2 infection or COVID-19 vaccination prior to the date of informed consent. Prior infection was defined as a positive test result for SARS-CoV-2 (either a reverse transcription-polymerase chain reaction test, other nucleic acid detection test, or SARS-CoV-2 antigen test), or a diagnosis of COVID-19.

Key exclusion criteria were the presence of serious cardiovascular, renal, hepatic, blood, neuropsychiatric or developmental disorder, thrombocytopenia, or coagulopathy; history of vaccine-related seizures or epilepsy, or history of anaphylaxis or severe allergy due to administration of food, drugs, cosmetics or vaccines; current or prior history of myocarditis or pericarditis; or a previous diagnosis of immunodeficiency or a close relative with congenital immunodeficiency. Persons who had tested positive for SARS-CoV-2 infection or were diagnosed with COVID-19within 3 months before the date of informed consent, or had symptoms suggestive of SARS-CoV-2 infection at the time of informed consent, or who had a positive SARS-COV-2 test at the time of eligibility evaluation were also ineligible for participation.

### Study design, treatments, and blinding

This was a phase 3 randomized, active-comparator, observer-blind, non-inferiority study registered with the Japan Registry of Clinical Trials with the identifier jRCT2031230424. The study was conducted in accordance with the Declaration of Helsinki, Good Clinical Practice guidelines, and all national and regional ordinance, and was approved by the relevant ethical committees at each study site.

The study design is shown in **Fig. 1A**. Participants were divided into subpopulations A–D according to their prior infection/vaccination status. The members of each subpopulation were then randomly assigned in a 1:1 ratio to receive either DS-5670d (60 µg of mRNA) or monovalent BNT162b2 (containing an antigen from omicron XBB.1.5, 30 µg of mRNA). The comparator BNT162b2 vaccine was supplied by the Japanese government to be used as the study control. Random assignment was the responsibility of an independent statistician, and vaccines were dispensed and administered intramuscularly into the deltoid region of the upper arm by unblinded site personnel on day 1. Investigators were blinded to treatment, and uploaded participant data via an interactive web response system. Participants and their legal representatives, the study monitor and members of the study advisory board, sponsor and collaborators, and personnel performing antibody titer determination were also blinded to vaccine assignment.

**Fig. 1.**
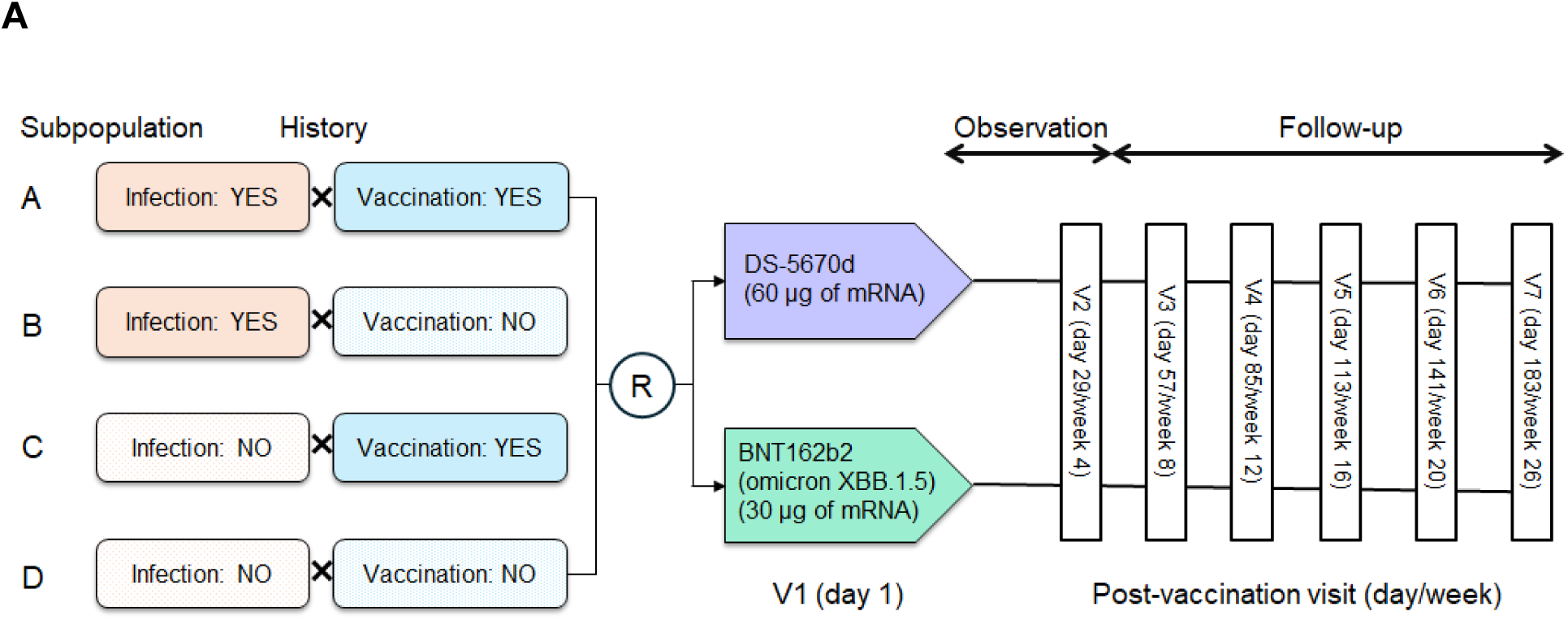
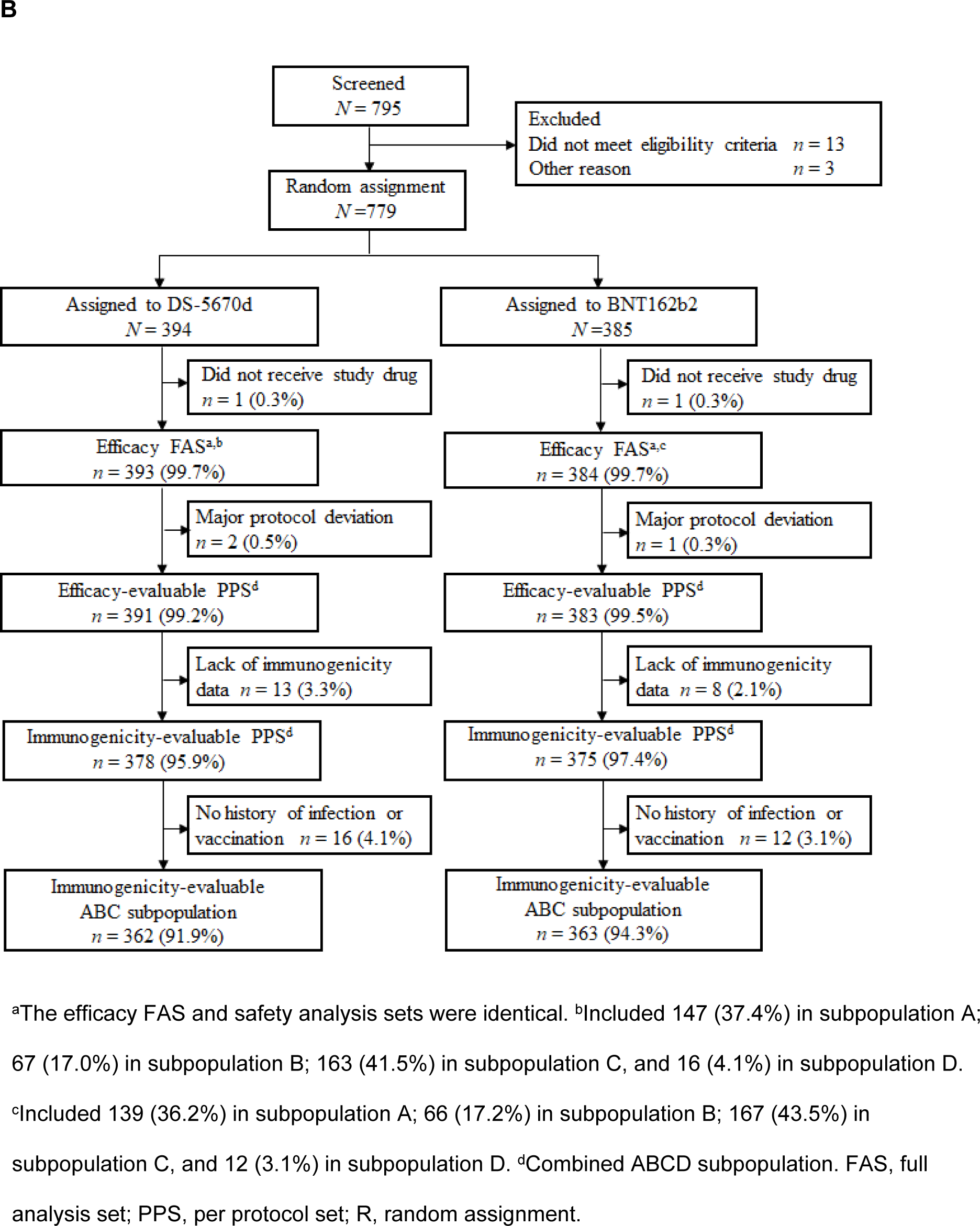
(A) Study design and (B) CONSORT flow diagram of participant disposition through the study.

The first participant was enrolled on 9 January 2024, the initial observation period lasted for 4 weeks post-vaccination, and the follow-up period was planned to continue up to 26 weeks post-vaccination. In the current manuscript, we report the results up to 4 weeks post-vaccination.

### Assessments and outcomes

The study objective was to verify the non-inferiority of DS-5670d to BNT162b2 in terms of immunogenicity, and to evaluate safety.

A sequential procedure was followed for the immunogenicity analyses. First, non-inferiority was tested for the combined ABC subpopulations; only if non-inferiority was confirmed was testing continued for the combined ABCD subpopulation. The primary efficacy endpoint was geometric mean titer (GMT) of blood neutralizing activity against SARS-CoV-2 (omicron variant XBB.1.5) and seroresponse rate at day 29 (4 weeks) after study vaccine administration in participants with any history of SARS-CoV-2 infection and/or vaccination (the combined ABC subpopulation). The key secondary efficacy endpoint was blood neutralizing activity GMT and seroresponse rate at day 29 after study vaccination in all study participants regardless of infection or vaccination history (the combined ABCD subpopulation). The incidence of COVID-19 (confirmed by the investigator based on a positive SARS-CoV-2 infection test plus any COVID-19 symptoms) up to day 29 after study vaccination was also evaluated.

Safety endpoints included the occurrence of solicited adverse events (AEs), unsolicited AEs, serious AEs (SAEs), and clinically relevant changes in laboratory values. AEs were coded using the Medical Dictionary for Regulatory Activities version 26.1. Health status was recorded in an electronic diary by participants or their representative. Solicited AEs (injection site events [redness, swelling, induration, pain, warmth, and pruritus] and systemic events [fever, malaise, headache, rash, and myalgia]) were collected up to day 8 post-administration, and unsolicited AEs and SAEs up to day 29. Laboratory tests were performed pre-vaccination and at day 29 post-vaccination.

### Statistical analysis

To confirm non-inferiority of DS-5670d versus BNT162b2 at day 29, the lower limit of the two-sided 95% confidence interval (CI) of the GMT ratio was required to be above the non-inferiority margin of 0.67, and the lower limit of the two-sided 95% CI for the difference in the seroresponse rate was required to exceed −10% in favor of DS-5670d. Based on this, plus taking into account anticipated dropout rates, the number of participants in the combined ABC subpopulations required to power the immunogenicity non-inferiority comparison was 690 (of whom 345 would receive DS-5670d and 345 would receive BNT162b2). Among these, 100 participants (50 per vaccine group) were required to be vaccine-naïve (i.e., included in subpopulation B). No sample size requirement or limitation was set for subpopulation D as the number of eligible participants for this group (no prior infection, no prior vaccination) was considered likely to be small.

Immunogenicity analyses were conducted based on the treatment group to which each participant was assigned; the primary analysis population was the immunogenicity-evaluable per protocol set (PPS), which included participants who received a dose of study drug, had a pre-administration and at least one post-administration immunogenicity measurement, and had no protocol violations that could affect the immunogenicity evaluations. Safety analyses were conducted based on the study drug that was actually administered. Unsolicited AEs were evaluated using the safety analysis set, which included all participants who received at least one dose of study drug; solicited AEs were evaluated in the solicited safety analysis set, which included all participants in the safety analysis set for whom information on the occurrence of at least one solicited AE was available.

Baseline participant characteristics were recorded descriptively as number (%), mean (standard deviation [SD]), or median (range). For the calculation of adjusted GMT (and two-sided 95% CI), a linear model was applied with common log transformed neutralizing titers as the dependent variable, the study vaccine group as the independent variable, and the common log transformed baseline titer and the categories of subpopulation as covariates. For the adjusted seroresponse rate, the between-group difference (and two-sided 95% CI) was calculated. The Mantel-Haenszel method was applied, and subpopulation was included as a stratum. The COVID-19 incidence rate was calculated using the PPS as the number of cases per 1000 person-years. Solicited AEs were recorded in an electronic diary by each participant or their legal representative; the presence or absence of solicited AEs and their severity were recorded up to 8 days post-vaccine administration, and any other (unsolicited) AEs were recorded up to day 29. The number (%) of participants with events were calculated according to the respective analysis set and tabulated. All statistical calculations were performed using SAS software version 9.4 or later (SAS Institute, Inc., Cary, NC, USA).

## Results

### Population

A total of 779 participants were enrolled into the study, of whom 394 were assigned to receive DS-5670d and 385 to receive BNT162b2 omicron XBB.1.5. The disposition and analysis populations are presented in **Fig. 1B**. One participant in each group did not receive study vaccination, so the safety analysis set included 393 participants in the DS-5670d group and 384 in the BNT162b2 group. Overall, the safety analysis set included 286 participants in subpopulation A, 133 in subpopulation B, 330 in subpopulation C, and 28 in subpopulation D.

After exclusions due to major protocol deviations and lack of immunogenicity data, the immunogenicity-evaluable PPS (combined subpopulations ABCD) included 378 participants in the DS-5670d group and 375 in the BNT162b2 group. For the primary efficacy endpoint, the 28 participants in subpopulation D were excluded; thus, the immunogenicity-evaluable ABC subpopulation comprised 362 in the DS-5670d group and 363 in the BNT162b2 group.

The demographics and baseline characteristics of participants are described in **Table 1**. The median age of participants was 46.0 years (range 12–90 years), with the majority aged between 18–65 years (644/777, 82.9%). Slightly more than half of participants were male (425/777, 54.7%), and had a history of SARS-CoV-2 infection (419/777, 53.9%). Many participants had received prior vaccination (616/777, 79.3%), with monovalent (original strain) vaccines in 313 (40.3%) of participants and bivalent (original strain/omicronBA.4-5) vaccines in 275 (35.4%). Characteristics were generally similar across the study vaccine groups, and there were no notable differences between the safety analysis set and the immunogenicity-evaluable PPS.

**Table 1.**
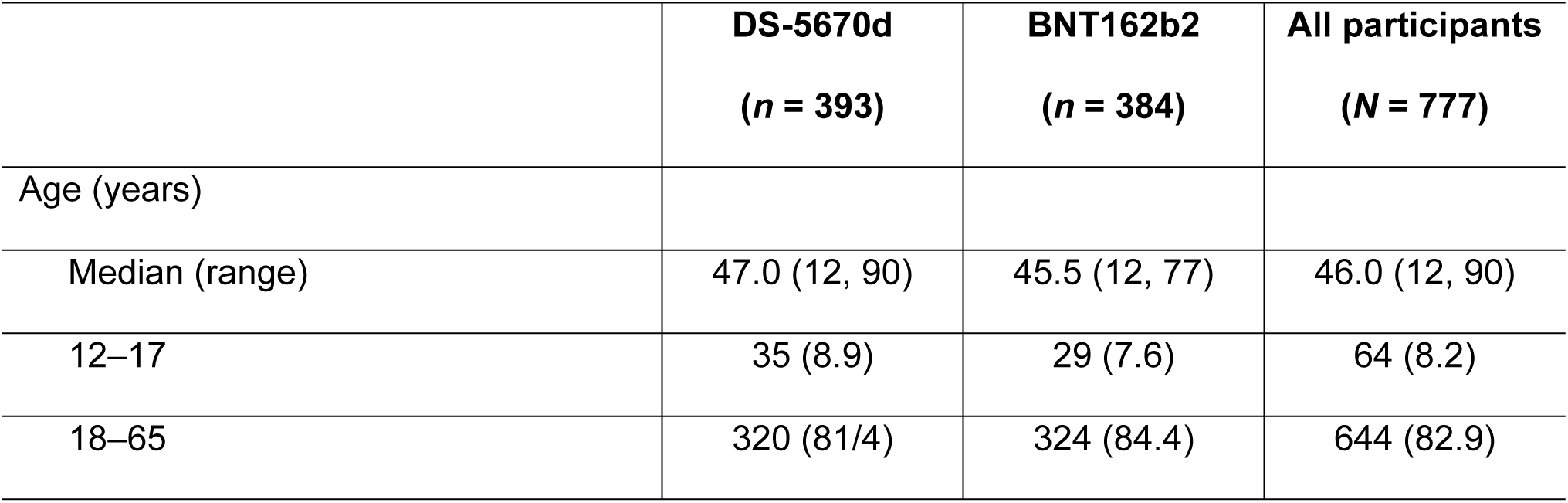
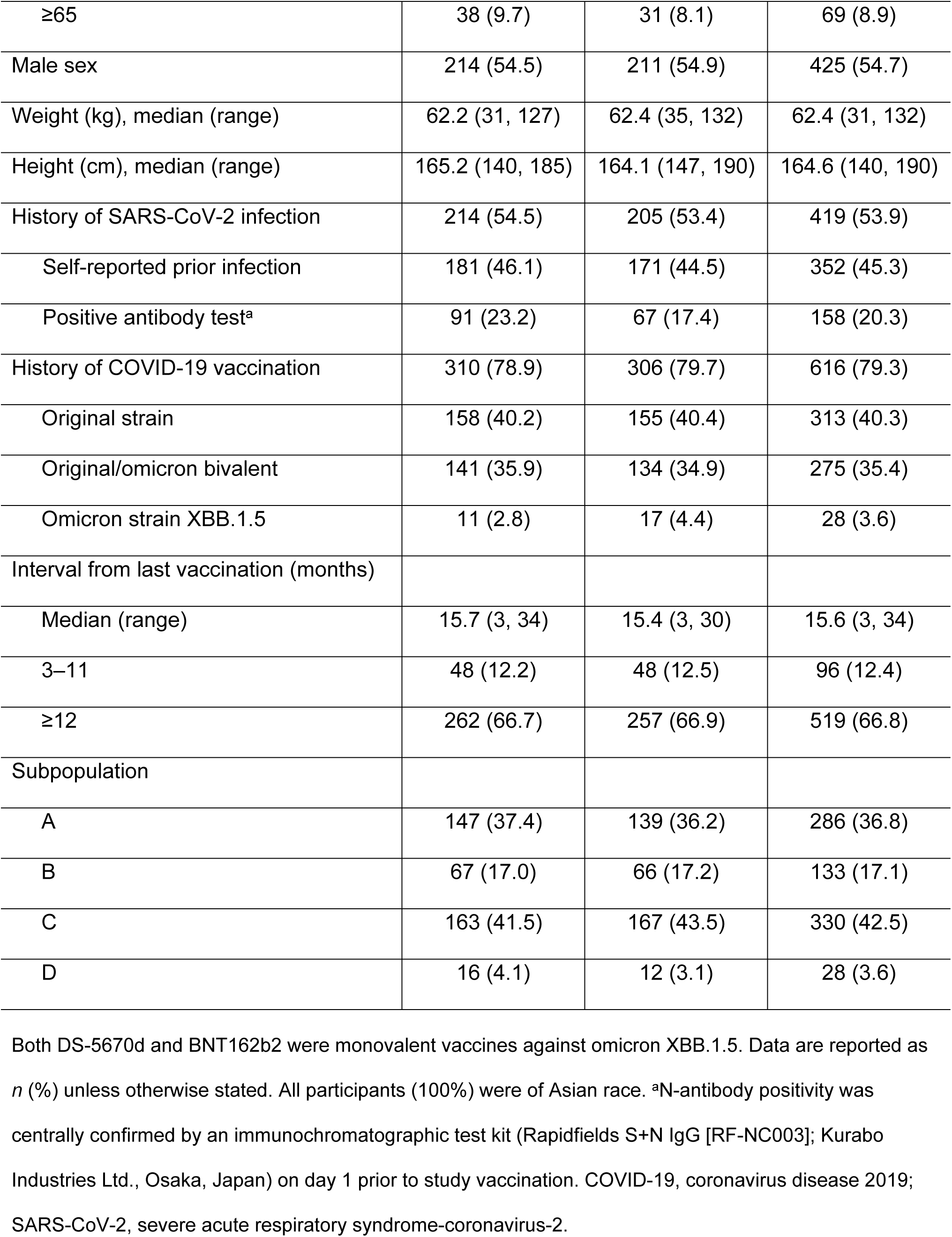
Baseline demographics and clinical characteristics (safety analysis set).

|  | **DS-5670d**<br>**(n = 393)** | **BNT162b2**<br>**(n = 384)** | **All participants**<br>**(N = 777)** |
| --- | --- | --- | --- |
| Age (years) |  |  |  |
| Median (range) | 47.0 (12, 90) | 45.5 (12, 77) | 46.0 (12, 90) |
| 12–17 | 35 (8.9) | 29 (7.6) | 64 (8.2) |
| 18–65 | 320 (81.4) | 324 (84.4) | 644 (82.9) |
| ≥65 | 38 (9.7) | 31 (8.1) | 69 (8.9) |
| Male sex | 214 (54.5) | 211 (54.9) | 425 (54.7) |
| Weight (kg), median (range) | 62.2 (31, 127) | 62.4 (35, 132) | 62.4 (31, 132) |
| Height (cm), median (range) | 165.2 (140, 185) | 164.1 (147, 190) | 164.6 (140, 190) |
| History of SARS-CoV-2 infection | 214 (54.5) | 205 (53.4) | 419 (53.9) |
| Self-reported prior infection | 181 (46.1) | 171 (44.5) | 352 (45.3) |
| Positive antibody test<sup>a</sup> | 91 (23.2) | 67 (17.4) | 158 (20.3) |
| History of COVID-19 vaccination | 310 (78.9) | 306 (79.7) | 616 (79.3) |
| Original strain | 158 (40.2) | 155 (40.4) | 313 (40.3) |
| Original/omicron bivalent | 141 (35.9) | 134 (34.9) | 275 (35.4) |
| Omicron strain XBB.1.5 | 11 (2.8) | 17 (4.4) | 28 (3.6) |
| Interval from last vaccination (months) |  |  |  |
| Median (range) | 15.7 (3, 34) | 15.4 (3, 30) | 15.6 (3, 34) |
| 3–11 | 48 (12.2) | 48 (12.5) | 96 (12.4) |
| ≥12 | 262 (66.7) | 257 (66.9) | 519 (66.8) |
| Subpopulation |  |  |  |
| A | 147 (37.4) | 139 (36.2) | 286 (36.8) |
| B | 67 (17.0) | 66 (17.2) | 133 (17.1) |
| C | 163 (41.5) | 167 (43.5) | 330 (42.5) |
| D | 16 (4.1) | 12 (3.1) | 28 (3.6) |

Both DS-5670d and BNT162b2 were monovalent vaccines against omicron XBB.1.5. Data are reported as *n* (%) unless otherwise stated. All participants (100%) were of Asian race. <sup>a</sup>N-antibody positivity was centrally confirmed by an immunochromatographic test kit (Rapidfields S+N IgG [RF-NC003]; Kurabo Industries Ltd., Osaka, Japan) on day 1 prior to study vaccination. COVID-19, coronavirus disease 2019; SARS-CoV-2, severe acute respiratory syndrome-coronavirus-2.

### Immunogenicity/efficacy

In the primary analysis (combined ABC subpopulations), the adjusted GMT ratio of DS-5670d to BNT162b2 was 1.218 (95% CI, 1.059, 1.401). Seroresponse rates were 87.3% in the DS-5670d group and 82.9% in the BNT162b2 group; the adjusted difference was 4.5% (95% CI, –0.70, 9.71). Both of these results exceeded the prespecified non-inferiority margins (**Table 2**), and the study met the primary endpoint.

**Table 2.**
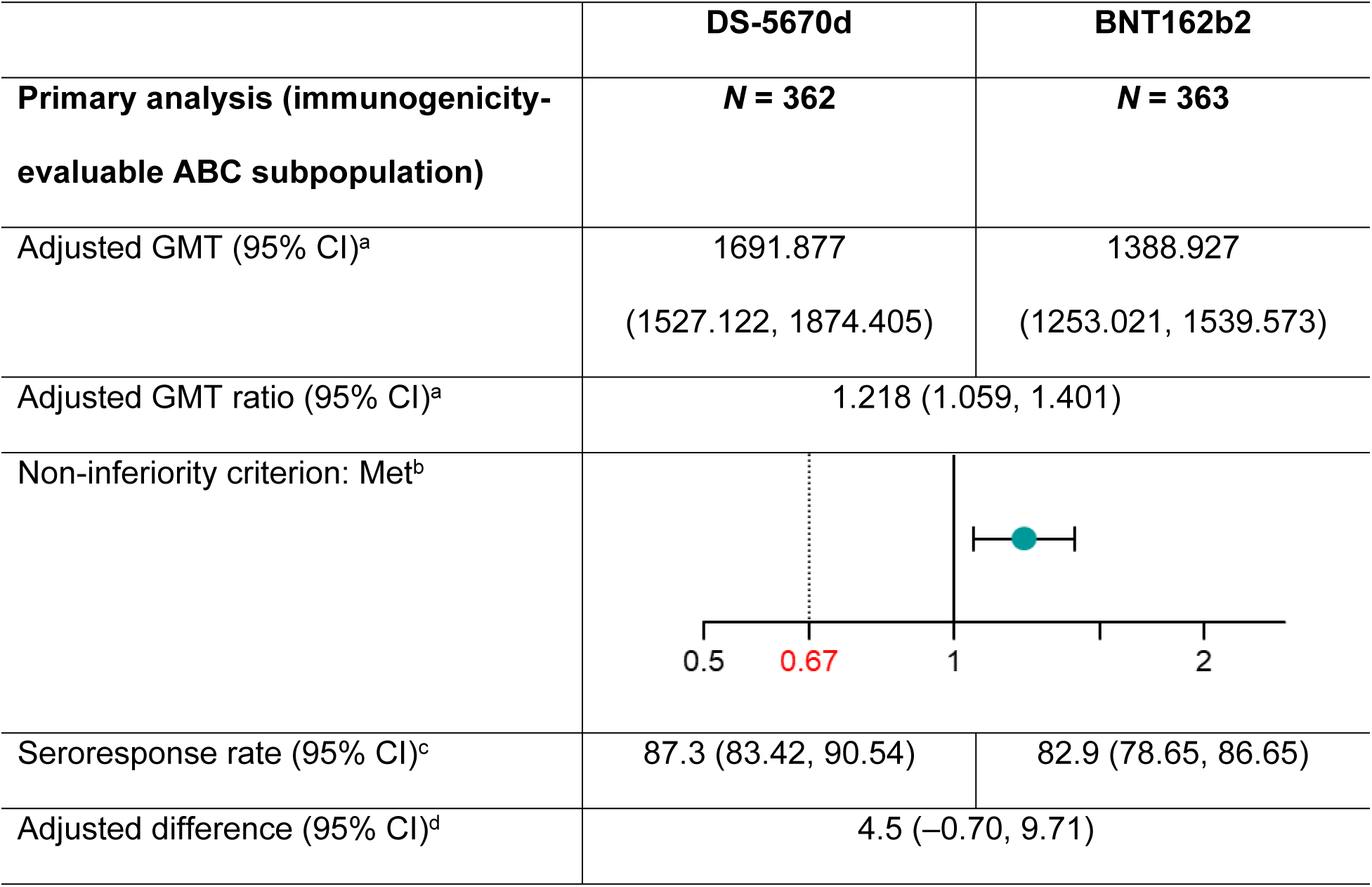
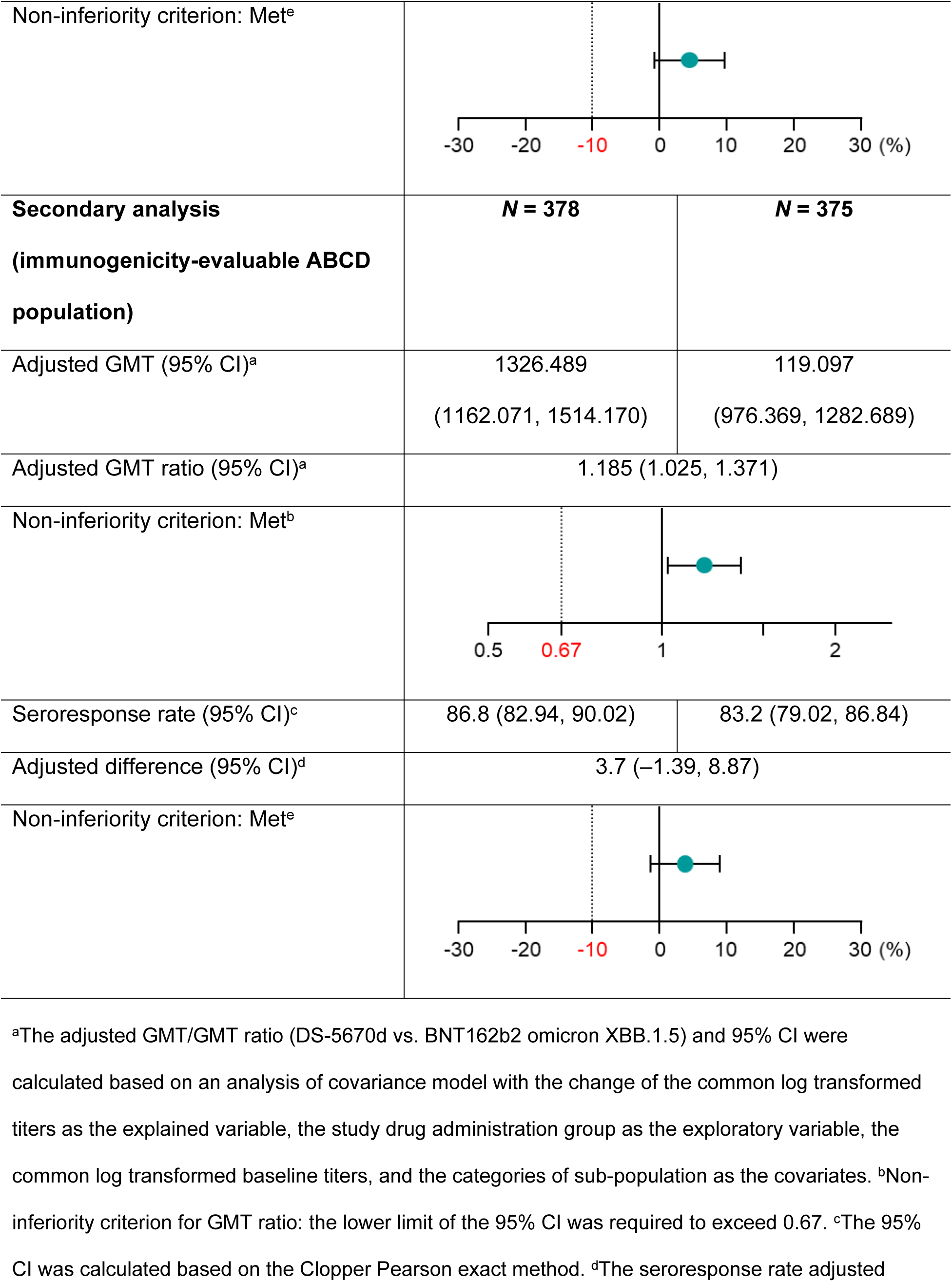
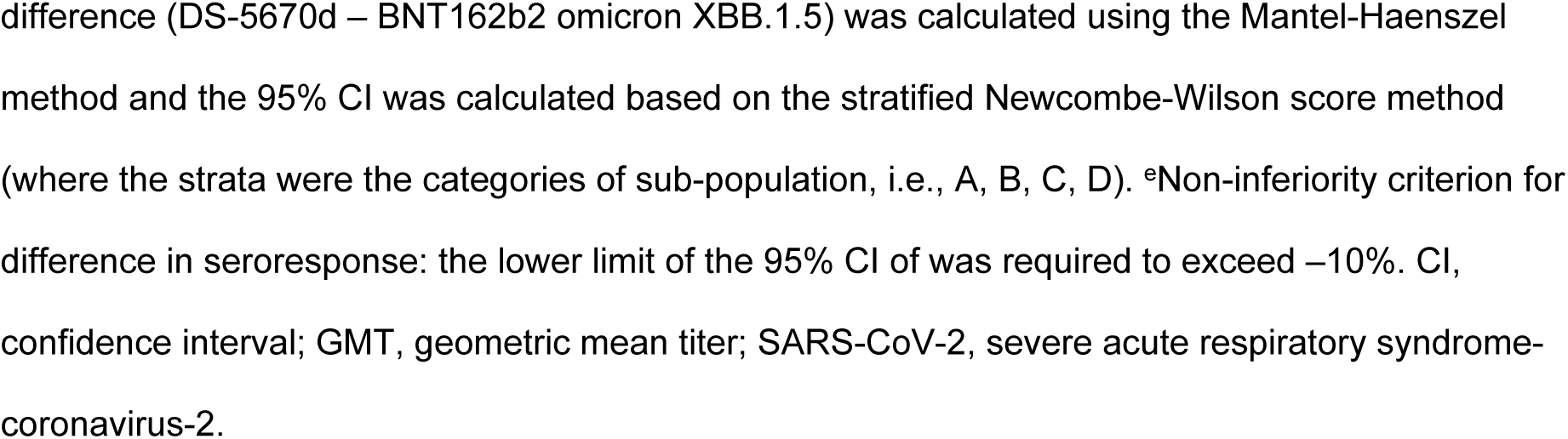
Comparison of neutralizing activities against SARS-CoV-2 (omicron XBB.1.5.6) at day 29.

In the secondary analysis (combined ABCD subpopulations), the adjusted GMT ratio was 1.185 (95% CI, 1.025, 1.371), and the adjusted difference in seroresponse rate was 3.7% (95% CI, –1.39, 8.87). Again, both results exceeded the prespecified non-inferiority margins (**Table 2**).

An analysis of blood neutralizing activity and seroresponse according to key subgroups (age group, sex, and subpopulation) is shown in **Fig. 2**. The GMT ratio tended to favor DS-5670d in all patients with a history of vaccination, and also in those with infection history but without a history of vaccination.

**Fig. 2.**
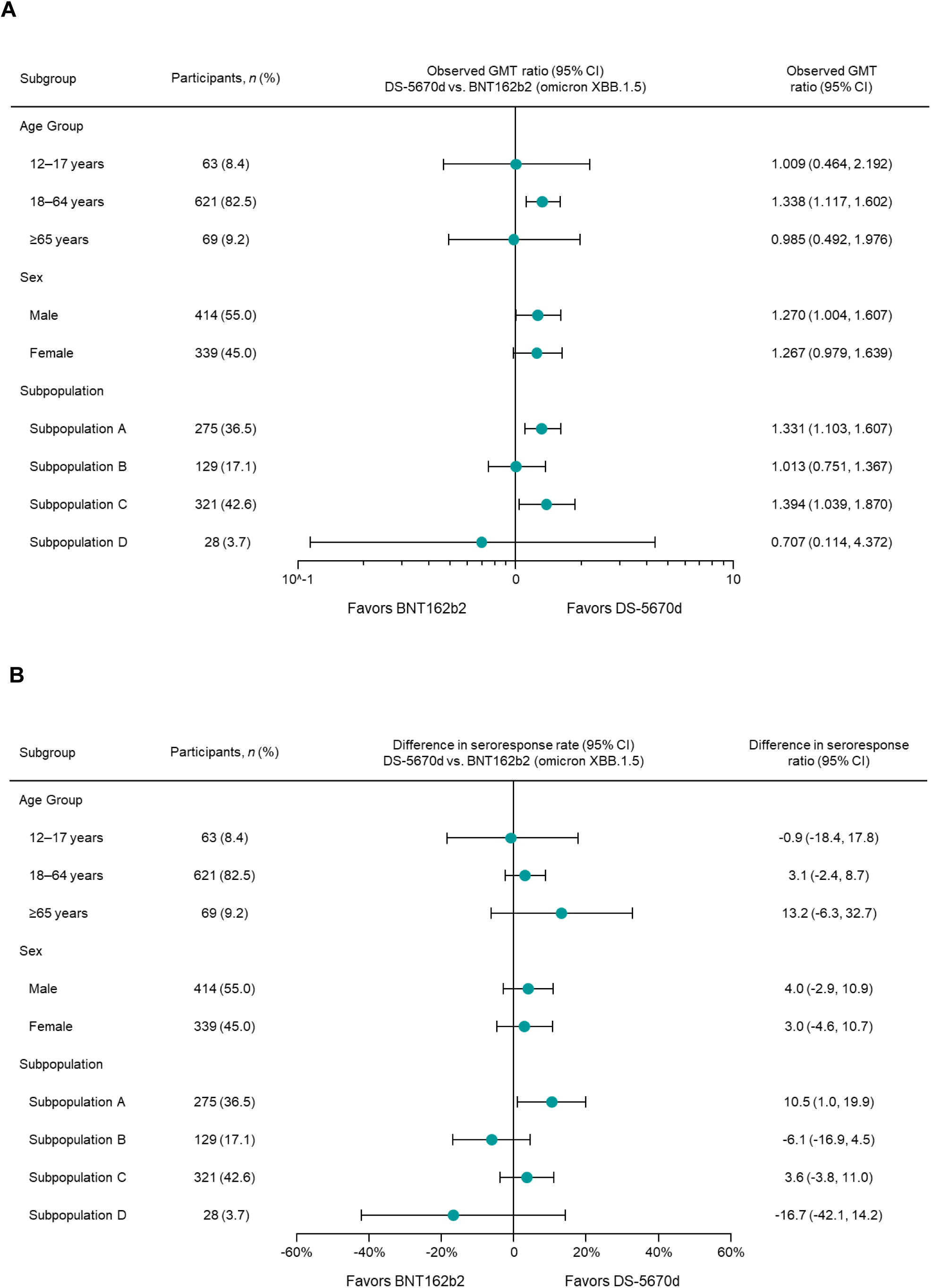
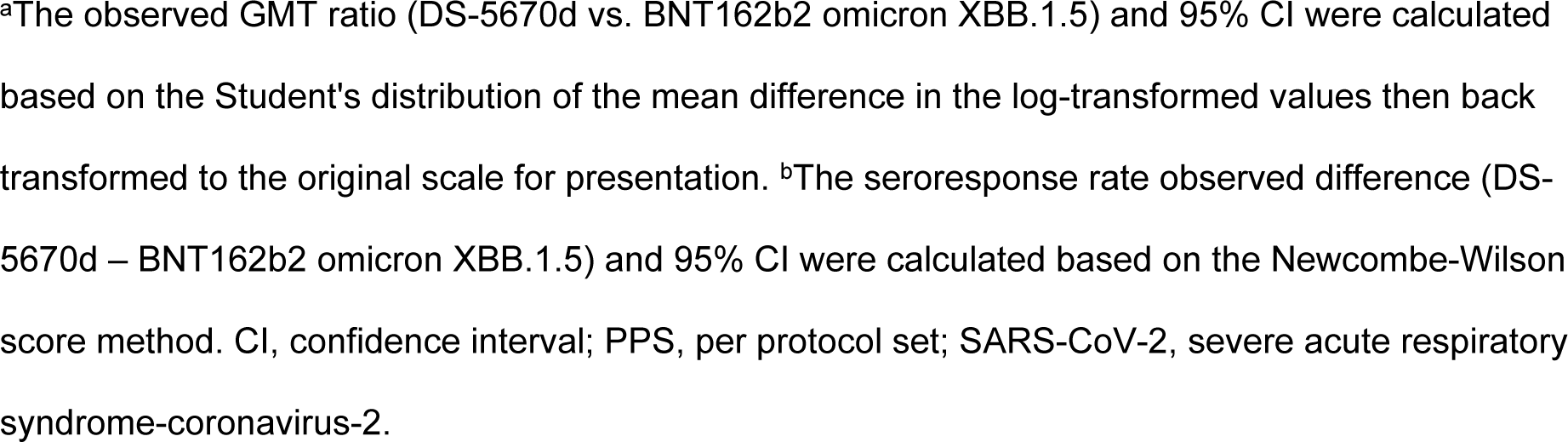
Forest plot of immunogenicity against SARS-CoV-2 (omicron XBB.1.5.6) at day 29 by subgroup for (A) observed GMT ratio of blood neutralizing activity^a^ and (B) seroresponse rate^b^ (immunogenicity-evaluable PPS).

The incidence rate of COVID-19 to day 29 after study vaccination is presented in **Fig. 3**. Both study vaccines appeared to be effective in preventing the onset of COVID-19 during this period, with two events reported in the DS-5670d group and three in the BNT162b2 group.

**Fig. 3.**
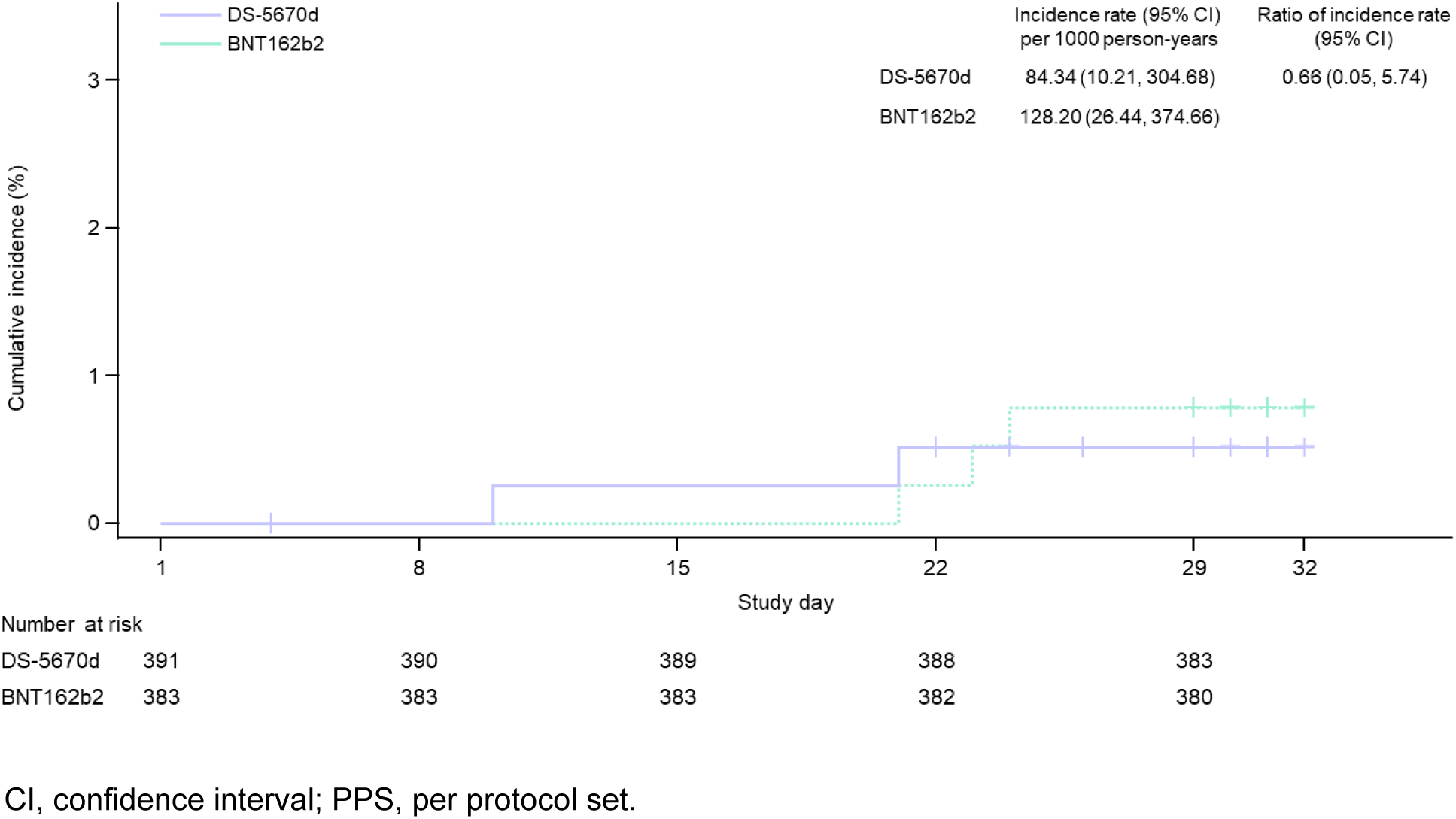
Incidence of investigator-confirmed COVID-19 (efficacy-evaluable PPS).

### Safety

AEs occurring during the study are summarized in **Table 3**. The majority of participants experienced at least one AE, with most AEs judged by the investigator to be vaccine-related. There were very few serious AEs across the study vaccine groups; none of the serious AEs were judged to be study vaccine-related.

**Table 3.** Summary of AEs (safety analysis set).

|  | **DS-5670d**<br>**(N = 393)** | **BNT162b2**<br>**(N = 384)** |
| --- | --- | --- |
| Participants with ≥1 solicited AE | 365 (92.9) | 348 (90.6) |
| Participants with ≥1 severe solicited AE | 20 (5.1) | 20 (5.2) |
| Participants with ≥1 solicited injection site AE | 361 (91.9) | 339 (88.3) |
| Participants with ≥1 severe solicited injection site AE | 14 (3.6) | 12 (3.1) |
| Participants with ≥1 solicited systemic AE | 183 (46.6) | 180 (46.9) |
| Participants with ≥1 severe solicited systemic AE | 7 (1.8) | 10 (2.6) |
| Participants with ≥1 unsolicited TEAE | 77 (19.6) | 74 (19.3) |
| Participants with ≥1 study vaccine-related unsolicited TEAE | 32 (8.1) | 14 (3.6) |
| Participants with ≥1 severe unsolicited TEAE | 0 | 0 |
| Participants with ≥1 serious TEAE | 0 | 2 (0.5) |
| Participants with ≥1 study vaccine-related serious TEAE | 0 | 0 |
| Participants with TEAE leading to discontinuation | 0 | 0 |
| Participants with fatal TEAE | 0 | 0 |

Data are shown as *n* (%). Solicited AEs were collected from the start of the study drug until day 8 after study vaccination. AE, adverse event; TEAE, treatment-emergent adverse event.

The majority of solicited AEs were mild or moderate in intensity (**Fig. 4**). The most frequently reported solicited injection site AE across both study vaccine groups was mild pain. Unsolicited treatment-emergent AEs (all causality and study vaccine-related) are described in **S2 Table**. Overall, there were no major differences in the incidence or severity of solicited or unsolicited AEs between the study vaccine groups.

**Fig. 4.**
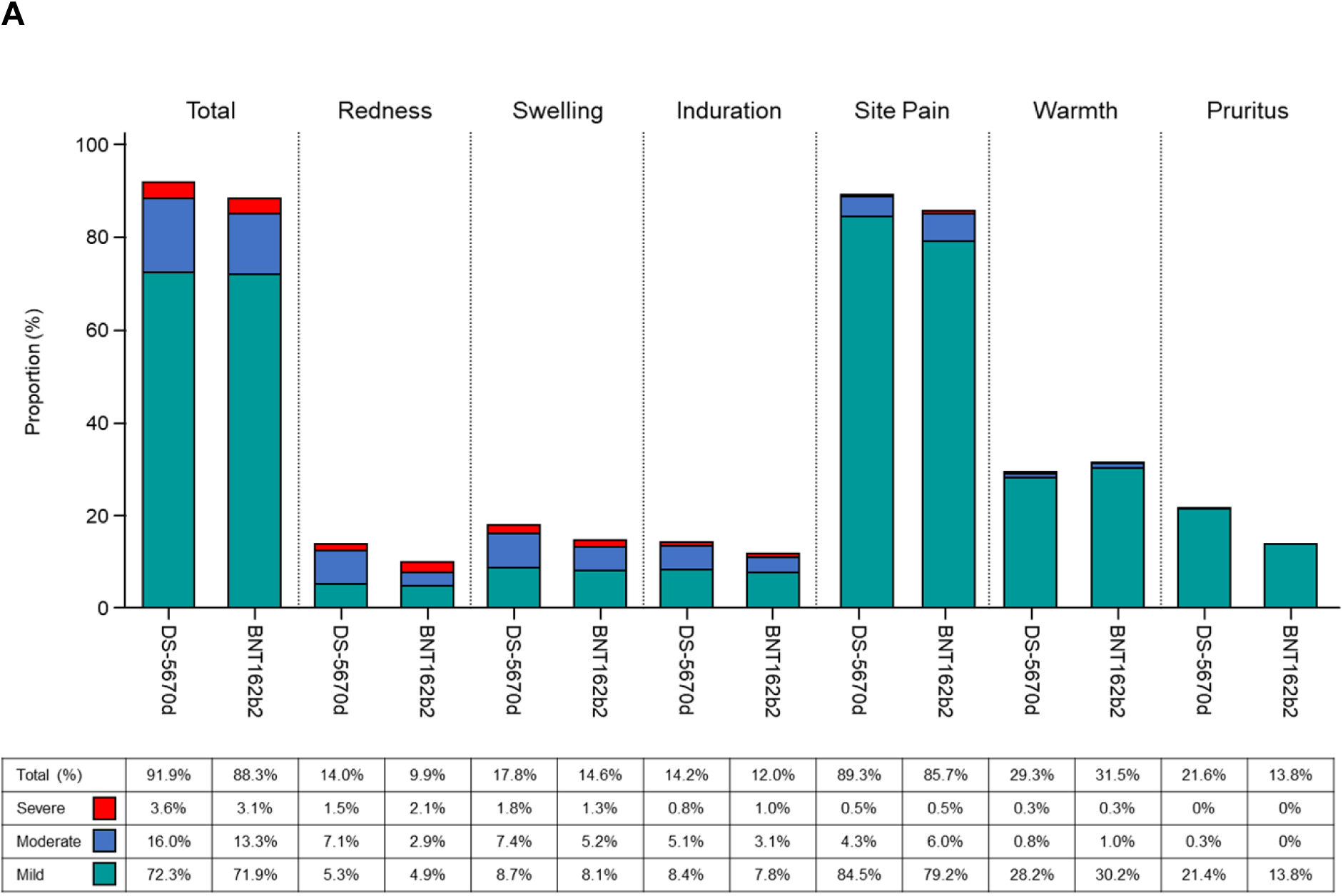
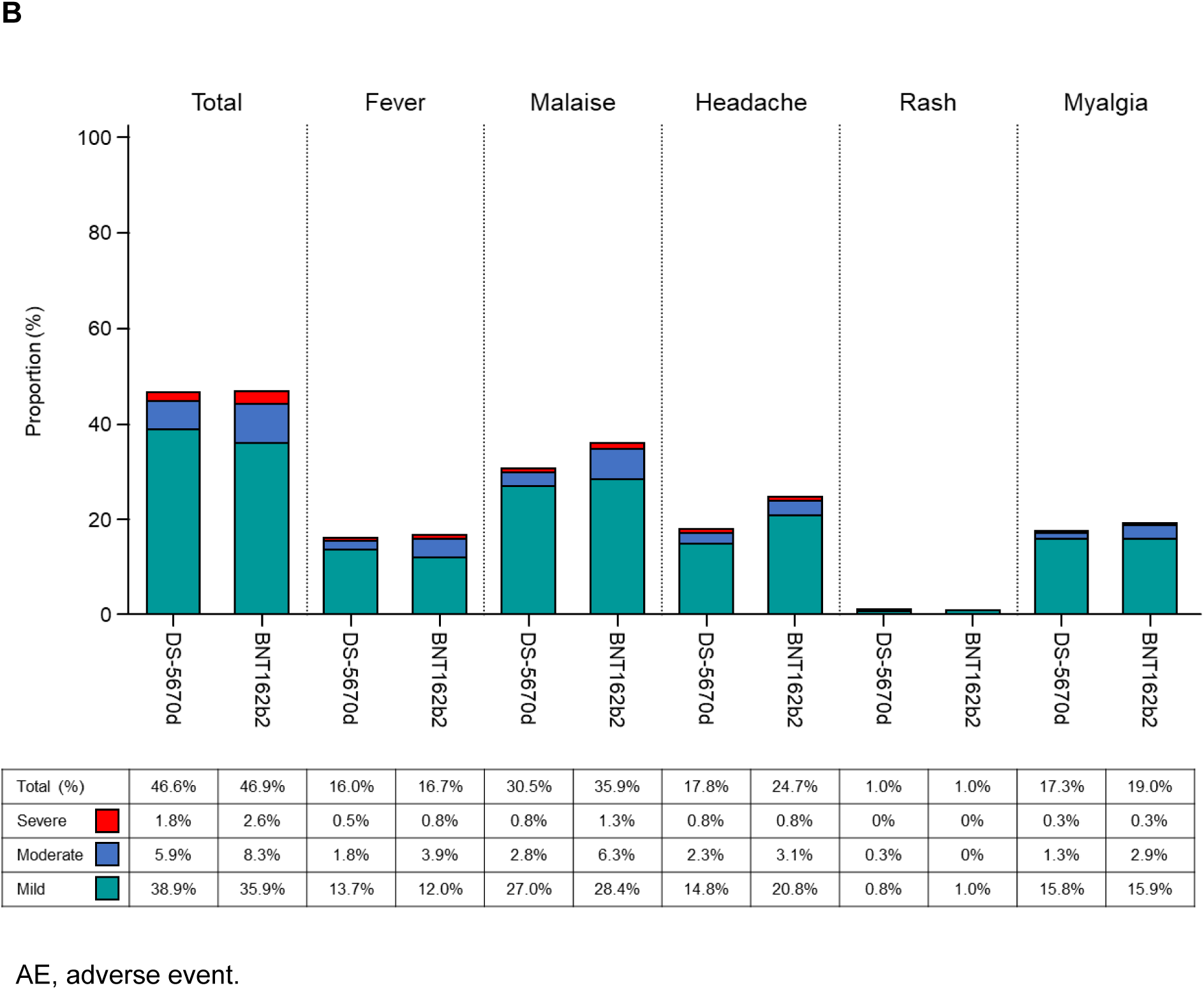
Solicited AEs (A) injection site AEs; (B) systemic AEs (safety analysis set).

## Discussion

In this study, non-inferior immunogenicity of a single dose of monovalent DS-5670d to BNT162b2 omicron XBB.1.5 was confirmed in participants aged ≥12 years, regardless of infection or vaccination history. Both the primary and secondary efficacy endpoints were met, with the non-inferiority criteria being met in both the combined ABC population (with any history of prior infection and/or vaccination), and in the overall PPS including participants without any history of prior infection or vaccination. In the combined ABC subpopulation, the adjusted GMT ratio (DS-5670d vs. BNT162b2) was 1.218 (95% CI, 1.059, 1.401), and the difference in the seroresponse rate was 4.5 (95% CI, –0.70, 9.71), both of which exceeded the non-inferiority margin.

In the analyses of immunogenicity by key demographic subgroups, there were no apparent differences in GMTs or seroresponse rates according to age group or sex. In addition to providing protective efficacy against severe COVID-19, infection-related death, and long COVID in high risk populations such as older adults [17, 18], recent evidence clearly indicates the benefits of COVID-19 vaccination for children and younger adults in terms of both their mental and physical health [19]. In a country such as Japan, where adults aged ≥65 comprise almost 30% of the population [20], continued vaccine coverage and infection prevention and control measure remain essential to protect at-risk and frail individuals, and sustain medical and social systems [21]. It is also important to achieve good immunogenicity in males, who typically have lower antibody responses to vaccination compared with females [22, 23], and which may impact the attainment of effective population immunity.

In the analyses of immunogenicity by subpopulation, the data suggested that even participants without any history of vaccination (subpopulation B) could achieve an adequate immune response with a single dose of DS-5670d. Such data emphasize the universality of DS-5670d immunogenicity regardless of prior vaccination history. In terms of participants without any history of infection or vaccination (subpopulation D), DS-5670d appears to be somewhat less immunogenic at first glance, but it is difficult to draw definitive conclusions due to the small number of participants available for evaluation. Overall, however, the participants enrolled in this study are a reasonably accurate reflection of the current real-world situation, where most people have now had a vaccination, or been infected, or both (i.e., comparable to the combined ABC population), and the number of those who have never been infected or vaccinated is dwindling. As such, the confirmation of non-inferiority of DS-5670d among both the ABC population and the ABCD population suggests that future seasonal DS-5670 vaccines will make a significant contribution to real-world public health in Japan.

The incidence of COVID-19 up to day 29 after study vaccination was low in both groups. However, the duration of observation was too short, and continued follow-up would be necessary before any inferences regarding the protective effects of DS-5670d can be made.

In terms of safety, there were no marked differences between the vaccine groups in the incidence or severity of solicited or unsolicited AEs. In line with results of previous studies evaluating different DS-5670 compositions, the most common solicited injection site AE was injection site pain, with most events being mild or moderate in severity [14, 15]. The most common solicited systemic AE in this study was malaise. This differed from the previously published data for DS-5670a in adults, where myalgia was the most common solicited systemic AE [14]. However, malaise was also the most common solicited systemic AE in studies of DS-5670a/b in both adults (Daiichi Sankyo Co., Ltd., data on file) and children [15]. Both injection site pain and malaise (or fatigue) are among the most frequently reported AEs among persons receiving mRNA vaccines against COVID-19, and are not unique to DS-5670 vaccines [24, 25]. There were no serious treatment-emergent unsolicited AEs which were judged to be related to study vaccine.

The main study limitation is the short duration of follow-up, which limits any conclusions relating to the long-term safety of DS-5670d, or the ongoing protection it may offer against COVID-19. In addition, the inclusion of only Asian patients may prevent generalization of the results to other locations and races.

In summary, the results of this phase 3 randomized, active-comparator, observer-blind study demonstrated the immunogenic non-inferiority of a single dose of DS-5670d regardless of history of prior infection or vaccination status and confirmed the clinically acceptable safety profile. Together with the data from previous DS-5670 clinical studies, these results contribute to the growing evidence that this LNP-mRNA vaccine platform technology can be harnessed to provide a significant contribution to public health via the production of COVID-19 vaccines in future seasons.

## Supporting Information

**S1 CONSORT Checklist.** Consolidated Standards of Reporting Trials (CONSORT) Checklist. (PDF)

**S1 Text. Study protocol.** (PDF)

**S1 Table. Listing of study sites and investigators.** (PDF)

**S2 Table. Unsolicited AEs occurring in >1 participant in either group (safety analysis set).** (PDF)

## Acknowledgments

The authors extend their thanks to all the participants and legal representatives who took part in the studies, and to the investigators and staff at the study sites for their support in conducting the study. The authors also thank all members of the clinical study team for valuable advice and support in the performance of this study. Finally, the authors thank Sally-Anne Mitchell, PhD, of McCann Health CMC, for providing medical writing support, which was funded by Daiichi Sankyo Co., Ltd.

## Data Availability Statement

The data underlying this report will be made available upon reasonable request.

## Abbreviations

AE: adverse event
COVID-19: coronavirus disease 2019
CI: confidence interval
GMT: geometric mean titer
LNP: lipid nanoparticle
mRNA: messenger ribonucleic acid
PPS: per protocol set
SAE: serious adverse event
SARS-CoV-2: severe acute respiratory syndrome-coronavirus-2
SD: standard deviation.

## References

1. Ulrichs T, Rolland M, Wu J, Nunes MC, El Guerche-Seblain C, Chit A. Changing epidemiology of COVID-19: Potential future impact on vaccines and vaccination strategies. Expert Rev Vaccines. 2024;23(1):510–22. doi: 10.1080/14760584.2024.2346589. PMID: 38656834.

2. Titball RW, Bernstein DI, Fanget NVJ, Hall RA, Longet S, MacAry PA, et al. Progress with COVID vaccine development and implementation. NPJ Vaccines. 2024;9(1):69. doi: 10.1038/s41541-024-00867-3. PMID: 38561358; PMCID: PMC10985065.

3. Dadras O, SeyedAlinaghi S, Karimi A, Shojaei A, Amiri A, Mahdiabadi S, et al. COVID-19 vaccines’ protection over time and the need for booster doses; A systematic review. Arch Acad Emerg Med. 2022;10(1):e53. doi: 10.22037/aaem.v10i1.1582. PMID: 36033989; PMCID: PMC9397599.

4. Teo SP. Review of COVID-19 mRNA vaccines: BNT162b2 and mRNA-1273. Journal of Pharmacy Practice. 2022;35(6):947–51. doi: 10.1177/08971900211009650. PMID: 33840294.

5. Goyal L, Zapata M, Ajmera K, Chaurasia P, Pandit R, Pandit T. A Hitchhiker’s Guide to worldwide COVID-19 vaccinations: A detailed review of monovalent and bivalent vaccine schedules, COVID-19 vaccine side effects, and effectiveness against omicron and delta variants. Cureus. 2022;14(10):e29837. doi: 10.7759/cureus.29837. PMID: 36204257; PMCID: PMC9527088.

6. Chatzilena A, Hyams C, Challen R, Marlow R, King J, Adegbite D, et al. Relative vaccine effectiveness of mRNA COVID-19 boosters in people aged at least 75 years during the spring-summer (monovalent vaccine) and autumn-winter (bivalent vaccine) booster campaigns: A prospective test negative case-control study, United Kingdom, 2022. Euro Surveill. 2023;28(48):2300173. doi: 10.2807/1560-7917.ES.2023.28.48.2300173. PMID: 38037728; PMCID: PMC10690860.

7. Murray CJL, Piot P. The potential future of the COVID-19 pandemic: Will SARS-CoV-2 become a recurrent seasonal infection? JAMA. 2021;325(13):1249–50. doi: 10.1001/jama.2021.2828. PMID: 33656519.

8. US Food and Drug Administration. Coronavirus (COVID-19) update: FDA authorizes changes to simplify use of bivalent mRNA COVID-19 vaccines; 18 April 2023. Available from: https://www.fda.gov/news-events/press-announcements/coronavirus-covid-19-update-fda-authorizes-changes-simplify-use-bivalent-mrna-covid-19-vaccines.

9. European Centre for Disease Prevention and Control, European Medicines Agency. ECDC-EMA statement on updating COVID-19 vaccines composition for new SARS-CoV-2 virus variants; 6 June 2023. Available from: https://www.ema.europa.eu/system/files/documents/other/ecdc-ema_statement_on_updating_covid-19_vaccines_composition_for_new_sars-cov-2_virus_variants_en.pdf.

10. Regan JJ, Moulia DL, Link-Gelles R, Godfrey M, Mak J, Najdowski M, et al. Use of updated COVID-19 vaccines 2023-2024 formula for persons aged ≥6 months: Recommendations of the Advisory Committee on Immunization Practices - United States, September 2023. MMWR Morb Mortal Wkly Rep. 2023;72(42):1140–6. doi: 10.15585/mmwr.mm7242e1. PMID: 37856366; PMCID: PMCPMC10602621.

11. European Medicines Agency. News: ETF recommends updating COVID-19 vaccines to target new JN.1 variant; 30 April 2024. Available from: https://www.ema.europa.eu/en/news/etf-recommends-updating-covid-19-vaccines-target-new-jn1-variant.

12. US Food and Drug Administration. Updated COVID-19 vaccines for use in the United States beginning in Fall 2024; 5 June 2024. Available from: https://www.fda.gov/vaccines-blood-biologics/updated-covid-19-vaccines-use-united-states-beginning-fall-2024.

13. Yabuta M, Takeshita F, Ueno S, Tanaka N, Yano T, Oe K, et al. Development of an mRNA vaccine against COVID-19. Trans Regulat Sci. 2021;3(3):118–9. doi: 10.33611/trs.2021-020.

14. Toyama K, Eto T, Takazawa K, Shimizu S, Nakayama T, Furihata K, et al. DS-5670a, a novel mRNA-encapsulated lipid nanoparticle vaccine against severe acute respiratory syndrome coronavirus 2: Results from a phase 2 clinical study. Vaccine. 2023;41(38):5525–34. doi: 10.1016/j.vaccine.2023.07.012. PMID: 37586958.

15. Suzuki R, Suda M, Ishida K, Furihata K, Ota A, Takahashii K, et al. Booster vaccination using bivalent DS-5670a/b is safe and immunogenic against SARS-CoV-2 variants in children aged 5-11 years: A phase 2/3, randomized, active-controlled study. Front Immunol. 2024;15:doi:10.3389/fimmu.2024.1445459. doi: 10.3389/fimmu.2024.1445459.

16. Daiichi-Sankyo. Press release: Daichirona® for intramuscular injection for booster vaccination approved in Japan as omicron XBB.1.5-adapted monovalent mRNA vaccine against COVID-19; 28 November 2023. Available from: https://www.daiichisankyo.com/files/news/pressrelease/pdf/202311/20231128_E.pdf.

17. Xu K, Wang Z, Qin M, Gao Y, Luo N, Xie W, et al. A systematic review and meta-analysis of the effectiveness and safety of COVID-19 vaccination in older adults. Front Immunol. 2023;14:1113156. doi: 10.3389/fimmu.2023.1113156. PMID: 36936964; PMCID: PMC10020204.

18. Mimura W, Ishiguro C, Terada-Hirashima J, Matsunaga N, Sato S, Kawazoe Y, et al. Effectiveness of BNT162b2 against infection, symptomatic infection, and hospitalization among older adults aged ≥65 years during the delta variant predominance in Japan: The VENUS study. J Epidemiol. 2024;34(6):278–85. doi: 10.2188/jea.JE20230106. PMID: 37743530; PMCID: PMC11078592.

19. McGrew S, Taylor HA. Adolescents, parents, and COVID-19 vaccination - Who should decide? N Engl J Med. 2022;386(2):e2. doi: 10.1056/NEJMp2116771. PMID: 34965336.

20. Japan Ministry of Internal Affairs and Communications. Statistics Topic No. 138: Japan’s elderly seen from statistics (Respect for the Aged Day); 18 September 2023. Available from: https://www.stat.go.jp/data/topics/pdf/topi138_01.pdf.

21. Hasegawa T, Hirata K, Matsumoto K. Lessons from the COVID-19 pandemic: Strategies and challenges for an aging society in Japan. Public Administration and Policy: An Asia-Pacific Journal. 2023;26(1):21–35. doi: 10.1108/PAP-10-2022-0123.

22. Flanagan KL, Fink AL, Plebanski M, Klein SL. Sex and gender differences in the outcomes of vaccination over the life course. Annu Rev Cell Dev Biol. 2017;33:577–99. doi: 10.1146/annurev-cellbio-100616-060718. PMID: 28992436.

23. Ciarambino T, Barbagelata E, Corbi G, Ambrosino I, Politi C, Lavalle F, et al. Gender differences in vaccine therapy: Where are we in COVID-19 pandemic? Monaldi Arch Chest Dis. 2021;91(4):1669. doi: 10.4081/monaldi.2021.1669. PMID: 33840183.

24. Kouhpayeh H, Ansari H. Adverse events following COVID-19 vaccination: A systematic review and meta-analysis. Int Immunopharmacol. 2022;109:108906. Epub 20220530. doi: 10.1016/j.intimp.2022.108906. PMID: 35671640; PMCID: PMC9148928.

25. Rabail R, Ahmed W, Ilyas M, Rajoka MSR, Hassoun A, Khalid AR, et al. The side effects and adverse clinical cases reported after COVID-19 immunization. Vaccines (Basel). 2022;10(4):488. doi: 10.3390/vaccines10040488. PMID: 35455237; PMCID: PMC9031559.

